# Circadian Patterns of Wearable-Derived Electrocardiographic Age and Left Atrial Remodeling in AF-Naïve Individuals

**DOI:** 10.64898/2026.03.17.26348661

**Authors:** Seung Hyun Park, Juhyun Jin, Jongwoo Kim, Dongha Lee, Daein Kim, Jaeseong Jang, Hee Tae Yu, Seng Chan You, Boyoung Joung

## Abstract

**Background:** AI-enabled electrocardiographic age (AI-ECG age) is a digital biomarker of electrophysiological cardiac health. Although cardiovascular physiology exhibits circadian organization, the circadian behavior of AI-ECG age and its structural correlates have not been defined in AF-naïve individuals.

**Objectives:** To determine whether AI-ECG age exhibits reproducible circadian patterns and whether disruption of these patterns is associated with left atrial (LA) remodeling, a marker of atrial myopathy.

**Methods:** Continuous single-lead wearable ECGs were analyzed from two independent prospective cohorts (S-Patch [https://ClinicalTrials.gov: NCT05119725, registered November 2021]; Memo Patch [https://ClinicalTrials.gov: NCT05355948, registered May 2022]). In AF-naïve participants with ≥48 hours of data, AI-ECG age was estimated every 10 minutes. Unsupervised clustering was used to identify intrinsic circadian trajectories. For clinical interpretability, participants were classified using a day–night difference cutoff (ΔAge 0.6 years) as Restorative (ΔAge >0.6) or Disrupted (ΔAge ≤0.6). We assessed phenotype reproducibility and examined associations with left atrial volume index (LAVI) using multivariable regression and meta-analysis.

**Results:** Unsupervised learning consistently identified three circadian trajectory patterns across cohorts. Under the simplified binary classification, the Restorative phenotype was observed in approximately half of the participants (47.6–50.2%). Phenotype reproducibility was moderate (Cohen’s κ=0.518; ICC=0.51–0.54) and was not fully explained by conventional heart rate variability measures. Among participants with echocardiography (n=122), the Disrupted phenotype was associated with higher LAVI (adjusted mean difference 6.09 mL/m²; 95% CI 1.46–10.72; p=0.010) and higher odds of severe LA enlargement (adjusted OR 4.17; 95% CI 1.58–10.99; p=0.004), with negligible heterogeneity (I²=0%).

**Conclusions:** Wearable-derived AI-ECG age exhibits circadian patterns in AF-naïve individuals, with unsupervised learning identifying distinct trajectories. Attenuation of a nocturnal decline—the Disrupted phenotype—is associated with left atrial enlargement, independent of conventional comorbidities and static AI-ECG age metrics. These findings suggest that circadian electrophysiological aging phenotyping may capture a dimension of atrial structural vulnerability not reflected by point-in-time assessments, and support prospective studies to evaluate its clinical utility.

**Condensed Abstract:** Continuous wearable ECGs revealed circadian patterns in AI-ECG age in AF-naïve individuals. A Disrupted phenotype, defined by an attenuated nocturnal decline (day–night AI-ECG age difference ≤0.6 years), was associated with left atrial remodeling, including higher LAVI and severe enlargement, independent of conventional comorbidities and static AI-ECG age metrics. Circadian electrophysiological aging phenotyping may capture a dimension of atrial structural vulnerability not reflected by point-in-time assessments, and motivates prospective validation of its clinical utility.

## Introduction

Physiological aging is a fundamental driver of cardiovascular disease and incident atrial fibrillation (AF) (1,2). Artificial intelligence (AI) has enabled the quantification of this biological process through “electrocardiographic age” (AI-ECG age)—a validated digital biomarker of electrophysiological health that robustly predicts mortality and cardiovascular outcomes (3–8). While AI-ECG age has traditionally been derived from static, hospital-based 12-lead ECGs, recent advances have translated this metric to ambulatory monitoring. We recently developed and validated the PROPHECG-Age Single model, which estimates AI-ECG age from continuous, single-lead wearable ECGs (9). This capability enables high-resolution circadian phenotyping of electrophysiological aging in free-living conditions, moving beyond episodic snapshots to characterize within-person temporal dynamics that may reveal underlying structural cardiac health.

Cardiovascular physiology is orchestrated by circadian biology and manifests as reproducible circadian variation in multiple clinical measures (10–12). Well-established day–night patterns in blood pressure dipping and heart rate variability (HRV) are strongly associated with clinical presentation and long-term cardiovascular prognosis (13–15). Recent evidence also suggests time-of-day patterning in AF episode onset and burden, including observations in screen-detected AF among individuals without prior clinically recognized AF (16), and contemporary reviews have highlighted circadian mechanisms that may shape AF susceptibility (17). However, whether AI-ECG age—a morphology-based marker of electrophysiological aging—exhibits reproducible circadian variation, and whether such patterns carry structural correlates, remains unclear.

To address this gap, we analyzed continuous single-lead wearable ECGs from two independent prospective registries (S-Patch and Memo Patch) in AF-naïve individuals. Because the circadian behavior of AI-ECG age has not been previously described, we first applied unsupervised machine learning to identify intrinsic circadian trajectories without imposing a priori assumptions. We then derived a clinically interpretable binary phenotype from these data-driven patterns, assessed its temporal reproducibility across consecutive days, and examined its association with adverse cardiac remodeling, quantified by the left atrial volume index (LAVI) (**Figure 1**).

**Figure 1.**
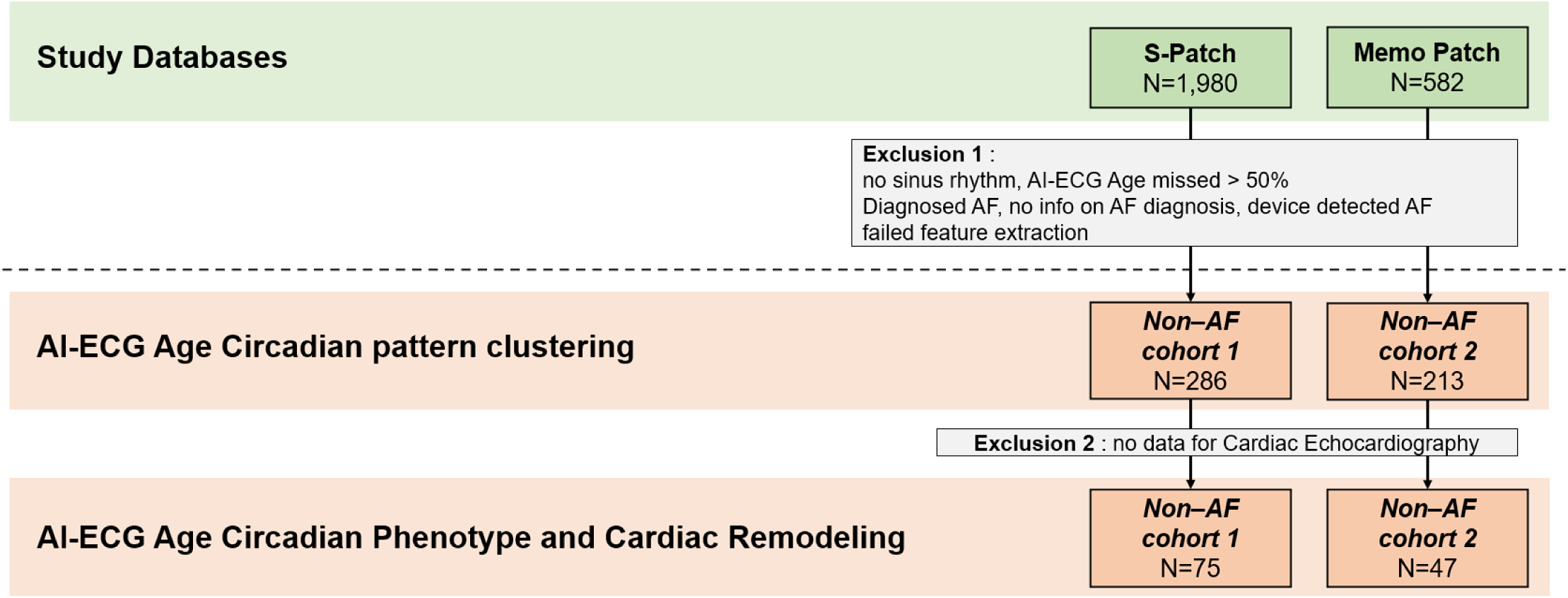
Overview of Study Design and Analysis Workflow. This figure illustrates the registry-based entry process and subsequent exclusions applied in the study. Participants were drawn from two wearable single-lead ECG databases: the S-Patch registry (N = 1,980) and the Memo-Patch registry (N = 582). Exclusion criteria included absence of sinus rhythm, >50% missing AI-ECG age data, diagnosed atrial fibrillation (AF), missing AF diagnostic information, device-detected AF, and failed feature extraction. After applying these exclusions, two AF-naive cohorts were derived for circadian pattern clustering: Cohort 1 (N = 286) from S-Patch and Cohort 2 (N = 213) from Memo-Patch. For analyses of AI-ECG age day–night gap and cardiac remodeling, only participants with available echocardiographic data were retained, yielding Cohort 1 (N = 75) and Cohort 2 (N = 47).

## Methods

### Study Design and Ethical Oversight

This study was performed in accordance with the ethical principles of the Declaration of Helsinki. For the prospective wearable validation cohorts, data were obtained from two pre-registered clinical trials approved by their respective Institutional Review Boards: The S-Patch registry (IRB No. 1-2021-0002; https://ClinicalTrials.gov identifier: NCT05119725, registered November 2021) and the Memo Patch registry (IRB No. 1-2022-0008; https://ClinicalTrials.gov identifier: NCT05355948, registered May 2022). Informed consent was obtained from all participants in these prospective registries.

### Study Population and Data

We analyzed continuous single-lead ECG data from two independent multicenter prospective registries, previously utilized to develop and validate the AI-ECG age estimation model in the PROPHECG-Age Single study (9). The S-Patch registry enrolled 1,980 adults across 15 centers in Korea (September 2021–August 2024), utilizing a chest-patch device for up to 72 hours of ambulatory monitoring. The Memo Patch registry included 582 adults from 13 centers (September 2022–November 2023), employing a patch-type device for extended monitoring of 7 to 14 days. To isolate physiological circadian phenotypes from arrhythmic variability, we applied strict exclusion criteria for the current analysis. Participants were excluded if they had (1) a prior clinical diagnosis of AF, (2) any AF episodes detected during the monitoring period, or (3) insufficient data quality or recording duration (<48 hours), as continuous monitoring covering at least two full circadian cycles was required to assess day–night consistency. For this analysis, AF-naïve was defined as no prior clinical diagnosis of AF and no AF detected during the index monitoring period.

### ECG Data Preprocessing and Feature Extraction

A standardized preprocessing pipeline was applied to ensure signal quality, utilizing the core methodology from our previously validated model (9). However, to minimize missing data and ensure robust temporal stability for circadian analysis, the sampling interval was optimized. Specifically, the highest-quality 10-second ECG epoch within each 10-minute window was extracted, yielding AI-ECG age estimates every 10 minutes over the first 48 hours of continuous monitoring. Following the exclusion of recordings with >50% missed epochs or failed feature extraction, 499 AF-naïve participants remained for the circadian analysis (S-Patch: n=286; Memo Patch: n=213).

Circadian features were derived from time- and frequency-domain representations to characterize AI-ECG age dynamics. AI-ECG age trajectories were mean-centered per individual before cosinor fitting; the cosinor model yielded amplitude, trough phase (bathyphase), and R² (18). A Fast Fourier Transform (FFT) identified the dominant frequency and corresponding period (“dominant_period”) (19). Day-to-day stability was quantified as self_similarity, defined as the Pearson correlation between the first and second 24-hour segments. Within-hour variability (“width”) was defined as the standard deviation of 10-minute AI-ECG age estimates within non-overlapping 60-minute windows, averaged across the 48-hour recording. The six features—dominant_period, width, self_similarity, cos_amplitude, cos_trough_phase, and cos_r2—were used for downstream dimensionality reduction and clustering.

### Circadian Phenotype Classification

All features were standardized (z-score) and transformed using kernel principal component analysis (RBF kernel, γ=0.2). K-means clustering was performed in the reduced feature space; the optimal number of clusters was selected by maximizing the average silhouette score across k=2–6.

For a clinically practical classification aligned with the clusters, we defined the day–night gap (ΔAge) as the difference between the mean daytime (10:00–16:00) AI-ECG ages and mean nighttime (00:00–06:00) AI-ECG ages (ΔAge = day_mean (AI-ECG age) − night_mean (AI-ECG age)), such that positive ΔAge indicates nocturnal dipping. Using a minimal-distance procedure on the S-Patch cohort, we identified an optimal threshold of ΔAge = 0.592 (sensitivity 0.581, specificity 0.667), which distinguished the Restorative (Rejuvenator) phenotype from the Disrupted (Riser + Irregular) phenotypes. For clinical simplicity, we adopted a rounded threshold of 0.6 years, which classified participants as Restorative (ΔAge > 0.6) and Disrupted (ΔAge ≤ 0.6). Applying the same threshold to the Memo Patch cohort yielded comparable operating characteristics (sensitivity 0.714, specificity 0.638), supporting a uniform cutoff across cohorts.

### Temporal Consistency of the Circadian Phenotype

We assessed temporal stability of the circadian AI-ECG age phenotype in 212 AF-naive participants from the Memo Patch registry with up to 7–14 days of continuous single-lead ECG monitoring. Each recording was partitioned into non-overlapping 48-hour epochs starting at the first available midnight anchor. For every epoch, we computed the day–night difference(ΔAge) and assigned the binary phenotype using the prespecified threshold: Restorative (ΔAge>0.6 years) or Disrupted (ΔAge≤0.6 years).

To evaluate the temporal stability of the binary circadian phenotype (Restorative=1, Disrupted=0), we performed a multi-step consistency analysis across sequential 48-hour epochs. First, for short-term stability, we compared the first two sequential epochs using a 2×2 contingency table, calculating percent agreement and Cohen’s κ. The association was further tested using χ^2^ with Cramer’s V for effect size, and Spearman’s ρ served as a monotonic agreement check. Second, for long-term consistency in participants with three (≈6 days) or four (≈8 days) epochs, we summarized across-epoch stability using percent agreement, Fleiss’ κ, and the mean of pairwise Cohen’s κ. Overall repeatability of the dichotomous labels was quantified using a two-way mixed-effects ICC (absolute agreement, single-measurement).

### Circadian Phenotype and Cardiac remodeling

The primary outcome was the left atrial volume index (LAVI, mL/m²) analyzed as a continuous variable; the secondary endpoint was severe left atrial enlargement defined as LAVI > 39 mL/m² (20). The key exposure was the ΔAge phenotype (Restorative: ΔAge > 0.6 years; Disrupted: ΔAge ≤ 0.6 years). Analyses were restricted to participants with echocardiography (n=122; S-Patch n=75; Memo Patch n=47). Within each cohort, associations between phenotype and continuous LAVI were estimated using ordinary least squares regression, first unadjusted and then adjusted for age, sex, BMI, and baseline AI-ECG age gap; effects are reported as mean differences (Disrupted − Restorative) with 95% CIs. For the binary endpoint, logistic regression with the same adjustment set was used, yielding odds ratios (Disrupted vs Restorative) and 95% CIs. Cohort-specific adjusted estimates and standard errors were then combined using inverse-variance meta-analysis; between-study heterogeneity was evaluated with Cochran’s Q, I², and τ² (REML estimator). When heterogeneity was negligible (I² ≈ 0), the fixed-effect summary served as the primary estimate.

### Secondary Analyses and Specificity Evaluation

To elucidate the underlying physiological mechanisms and evaluate the clinical specificity of the circadian AI-ECG age phenotype, we performed several prespecified secondary analyses. First, autonomic nervous system integrity was assessed using standard time-domain heart rate variability (HRV) indices, including the standard deviation of normal-to-normal intervals (SDNN), the root mean square of successive differences (RMSSD), and the percentage of adjacent NN intervals differing by >50 ms (pNN50). To confirm that the “Disrupted” phenotype was an independent physiological trait rather than a byproduct of systemic disease or general cardiac aging, we further scrutinized its associations with traditional cardiovascular comorbidities—hypertension, diabetes, and heart failure—as well as the static mean AI-ECG age. Additionally, a comparative analysis was performed to contrast the pathophysiological implications of the static AI-ECG age gap versus the circadian phenotype by evaluating their respective associations with left ventricular ejection fraction (LVEF) and LAVI. Finally, all primary and secondary associations were stratified by sex to identify potential sexual dimorphism in the manifestation and structural correlates of circadian electrophysiological disruption.

### Statistical Analysis

Continuous variables are presented as mean ± standard deviations and categorical variables as counts (percentages). Group comparisons used Welch’s t-test or the Mann–Whitney U test for continuous variables and χ² or Fisher’s exact tests for categorical variables. Due to the skewed distribution of HRV metrics, these variables were log-transformed prior to statistical analysis to satisfy normality assumptions. Two-sided p<0.05 was considered statistically significant. Analyses were performed in Python (NumPy, pandas, SciPy) and R (v4.4.3), including the meta package (v6.5-0).

### Data Availability

Anonymized data used in this study will be made available to qualified investigators for the purpose of replicating the analyses and findings, subject to appropriate ethical approvals and institutional authorizations.

## Results

### Baseline Characteristics and Cohort Heterogeneity

The study population comprised 499 participants across two independent cohorts with distinct clinical profiles (**Table 1**). The Memo Patch cohort (n=213) represented an older population (67.0 ± 9.7 years vs. 63.0 ± 13.8 years; p <0.001) with a higher proportion of female participants (76.1% vs. 51.4%; p <0.001) and a greater burden of comorbidities, as indicated by higher CHA₂DS₂-VASc scores (p <0.001), compared with the S-Patch cohort (n=286). Structurally, the Memo Patch group exhibited more advanced left atrial remodeling than the S-Patch group, characterized by a higher left atrial volume index (LAVI: 37.46 ± 15.48 vs. 31.35 ± 11.77 mL/m²; p = 0.023) and larger left atrial anteroposterior diameter (38.22 ± 6.42 vs. 35.41 ± 6.22 mm; p = 0.015). Despite being chronologically older, the Memo Patch cohort showed comparable AI-predicted heart ages to the S-Patch cohort, resulting in a lower AI-ECG age gap (−7.48 years vs. −3.77 years; p <0.001).

**Table 1.**
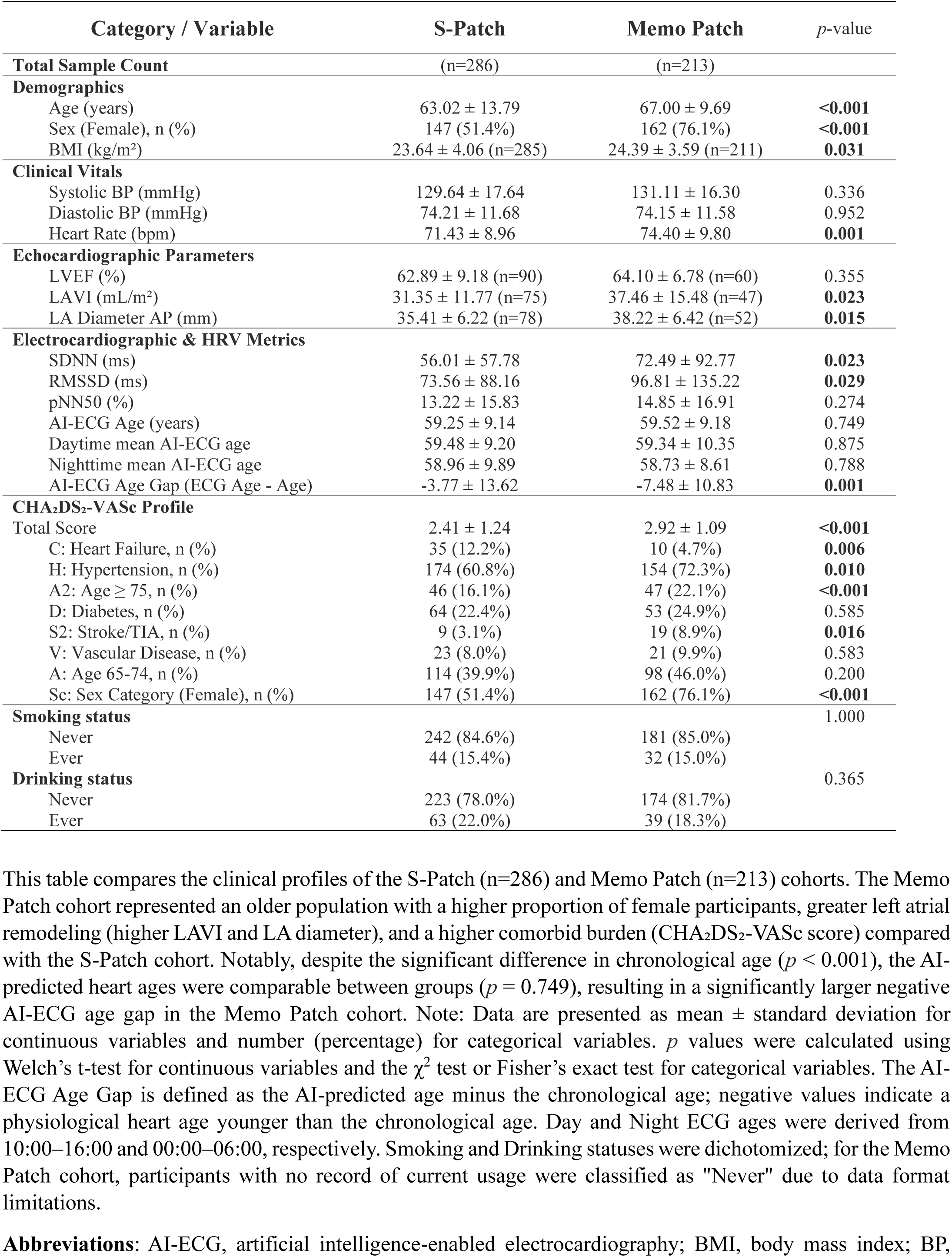
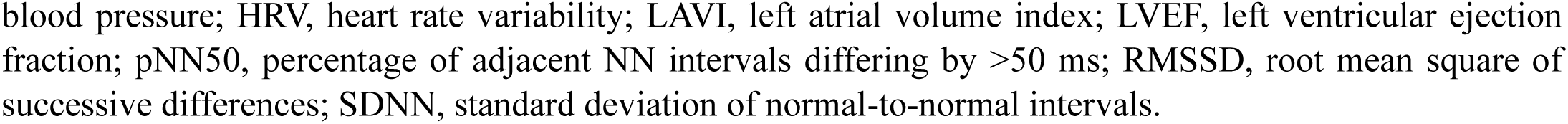
Baseline Clinical, Echocardiographic, and AI-Derived Electrocardiographic Characteristics by Study Cohort.

| Category / Variable | S-Patch | Memo Patch | *p*-value |
| --- | --- | --- | --- |
| **Total Sample Count** | (n=286) | (n=213) |  |
| **Demographics** |  |  |  |
| Age (years) | 63.02 ± 13.79 | 67.00 ± 9.69 | **<0.001** |
| Sex (Female), n (%) | 147 (51.4%) | 162 (76.1%) | **<0.001** |
| BMI (kg/m<sup>2</sup>) | 23.64 ± 4.06 (n=285) | 24.39 ± 3.59 (n=211) | **0.031** |
| **Clinical Vitals** |  |  |  |
| Systolic BP (mmHg) | 129.64 ± 17.64 | 131.11 ± 16.30 | 0.336 |
| Diastolic BP (mmHg) | 74.21 ± 11.68 | 74.15 ± 11.58 | 0.952 |
| Heart Rate (bpm) | 71.43 ± 8.96 | 74.40 ± 9.80 | **0.001** |
| **Echocardiographic Parameters** |  |  |  |
| LVEF (%) | 62.89 ± 9.18 (n=90) | 64.10 ± 6.78 (n=60) | 0.355 |
| LAVI (mL/m<sup>2</sup>) | 31.35 ± 11.77 (n=75) | 37.46 ± 15.48 (n=47) | **0.023** |
| LA Diameter AP (mm) | 35.41 ± 6.22 (n=78) | 38.22 ± 6.42 (n=52) | **0.015** |
| **Electrocardiographic & HRV Metrics** |  |  |  |
| SDNN (ms) | 56.01 ± 57.78 | 72.49 ± 92.77 | **0.023** |
| RMSSD (ms) | 73.56 ± 88.16 | 96.81 ± 135.22 | **0.029** |
| pNN50 (%) | 13.22 ± 15.83 | 14.85 ± 16.91 | 0.274 |
| AI-ECG Age (years) | 59.25 ± 9.14 | 59.52 ± 9.18 | 0.749 |
| Daytime mean AI-ECG age | 59.48 ± 9.20 | 59.34 ± 10.35 | 0.875 |
| Nighttime mean AI-ECG age | 58.96 ± 9.89 | 58.73 ± 8.61 | 0.788 |
| AI-ECG Age Gap (ECG Age - Age) | -3.77 ± 13.62 | -7.48 ± 10.83 | **0.001** |
| **CHA<sub>2</sub>DS<sub>2</sub>-VASc Profile** |  |  |  |
| Total Score | 2.41 ± 1.24 | 2.92 ± 1.09 | **<0.001** |
| C: Heart Failure, n (%) | 35 (12.2%) | 10 (4.7%) | **0.006** |
| H: Hypertension, n (%) | 174 (60.8%) | 154 (72.3%) | **0.010** |
| A2: Age ≥ 75, n (%) | 46 (16.1%) | 47 (22.1%) | **<0.001** |
| D: Diabetes, n (%) | 64 (22.4%) | 53 (24.9%) | 0.585 |
| S2: Stroke/TIA, n (%) | 9 (3.1%) | 19 (8.9%) | **0.016** |
| V: Vascular Disease, n (%) | 23 (8.0%) | 21 (9.9%) | 0.583 |
| A: Age 65-74, n (%) | 114 (39.9%) | 98 (46.0%) | 0.200 |
| Sc: Sex Category (Female), n (%) | 147 (51.4%) | 162 (76.1%) | **<0.001** |
| **Smoking status** |  |  | 1.000 |
| Never | 242 (84.6%) | 181 (85.0%) |  |
| Ever | 44 (15.4%) | 32 (15.0%) |  |
| **Drinking status** |  |  | 0.365 |
| Never | 223 (78.0%) | 174 (81.7%) |  |
| Ever | 63 (22.0%) | 39 (18.3%) |  |

This table compares the clinical profiles of the S-Patch (n=286) and Memo Patch (n=213) cohorts. The Memo Patch cohort represented an older population with a higher proportion of female participants, greater left atrial remodeling (higher LAVI and LA diameter), and a higher comorbid burden (CHA<sub>2</sub>DS<sub>2</sub>-VASc score) compared with the S-Patch cohort. Notably, despite the significant difference in chronological age ( $p < 0.001$ ), the AI-predicted heart ages were comparable between groups ( $p = 0.749$ ), resulting in a significantly larger negative AI-ECG age gap in the Memo Patch cohort. Note: Data are presented as mean ± standard deviation for continuous variables and number (percentage) for categorical variables. $p$ values were calculated using Welch's t-test for continuous variables and the $\chi^2$ test or Fisher's exact test for categorical variables. The AI-ECG Age Gap is defined as the AI-predicted age minus the chronological age; negative values indicate a physiological heart age younger than the chronological age. Day and Night ECG ages were derived from 10:00–16:00 and 00:00–06:00, respectively. Smoking and Drinking statuses were dichotomized; for the Memo Patch cohort, participants with no record of current usage were classified as "Never" due to data format limitations.
**Abbreviations:** AI-ECG, artificial intelligence-enabled electrocardiography; BMI, body mass index; BP,
blood pressure; HRV, heart rate variability; LAVI, left atrial volume index; LVEF, left ventricular ejection fraction; pNN50, percentage of adjacent NN intervals differing by >50 ms; RMSSD, root mean square of successive differences; SDNN, standard deviation of normal-to-normal intervals.

In sex-specific analyses (**Table S1**), AI-ECG age metrics were generally similar between sexes, whereas structural and lifestyle differences were observed. In the S-Patch cohort, male participants had larger LA diameters than females (37.29 ± 7.07 vs. 33.43 ± 4.46 mm; p = 0.005). Conversely, in the Memo Patch cohort, female participants exhibited higher LAVI compared to males (39.95 ± 16.35 vs. 30.22 ± 9.93 mL/m²; p = 0.020). Lifestyle factors such as smoking and alcohol consumption were more prevalent in male participants across both cohorts (p <0.001 for all). These heterogeneous demographic and physiological baselines provided a framework for validating the circadian AI-ECG phenotype across distinct clinical settings.

### Identification of Circadian AI-ECG Age Patterns

Unsupervised k-means clustering of 48-hour AI-ECG age trajectories revealed three consistent patterns in both registries (**Figure S1**): a Rejuvenator (R) pattern with a pronounced nocturnal decline, a Riser (Ri) pattern with a blunted or paradoxical nocturnal increase, and an Irregular (I) pattern with minimal rhythmicity. The optimal cluster number was k=3 for each dataset (S-Patch: silhouette=0.513; Memo Patch: silhouette=0.470). These circadian patterns were concordant across registries, with R-types reaching their nadir near midnight (trough time: 0.04 h in S-Patch; 1.14 h in Memo Patch; **Figure 2A-B**), Ri-types in the afternoon (14.64 h in S-Patch; 14.22 h in Memo Patch; **Figure 2C-D**), whereas I-types lacked a coherent circadian structure (**Figure 2E-H**).

**Figure 2.**
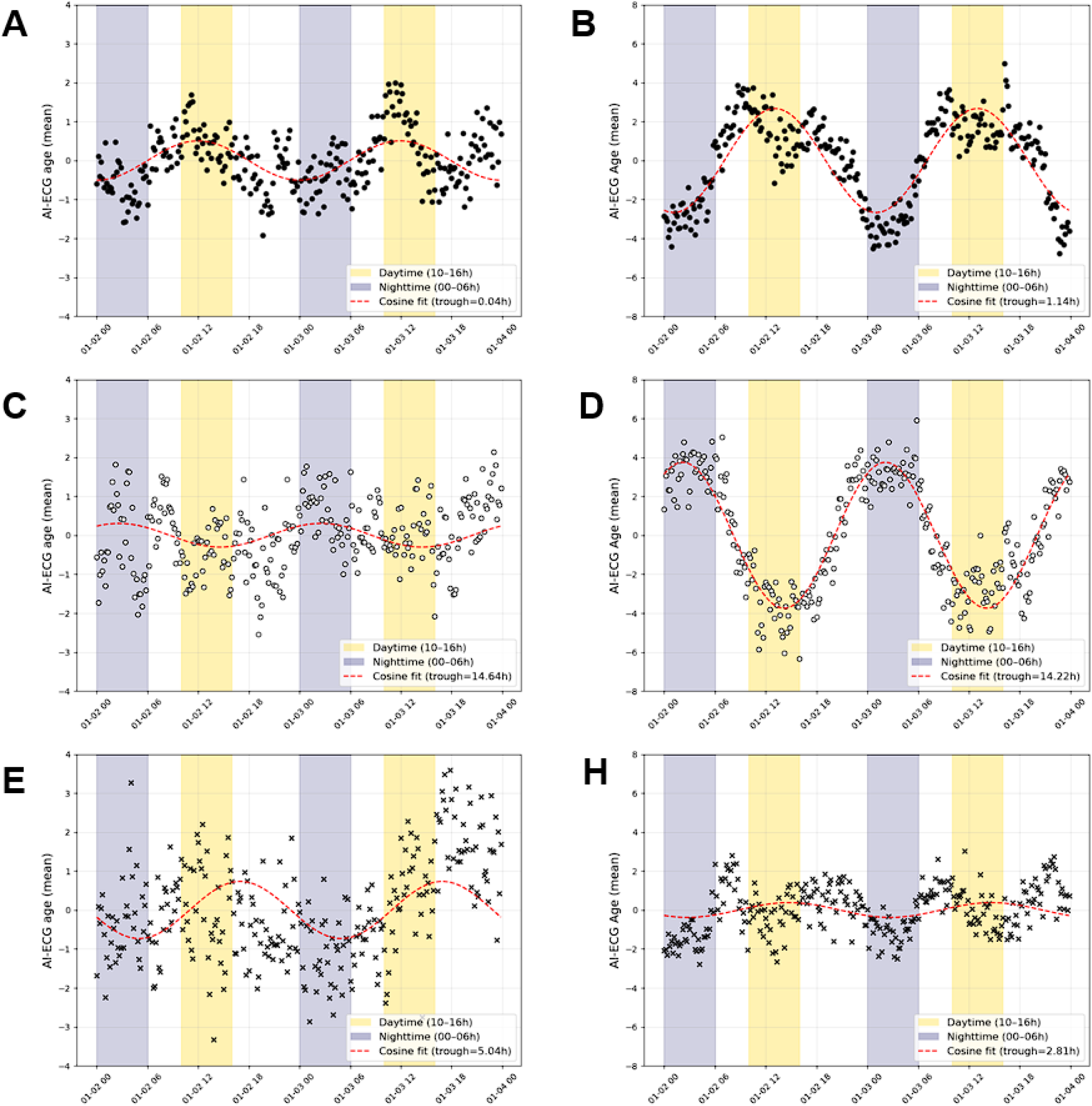
Circadian Clustering of 48-Hour AI-ECG Age Trajectories. This figure shows representative circadian patterns of AI-ECG age trajectories over 48 hours in atrial fibrillation (AF)-naive participants from the S-Patch (Cohort 1) and Memo-Patch (Cohort 2) registries. Participants with sinus rhythm and ≥50% valid AI-ECG age estimates were included, while those with diagnosed or device-detected AF, missing AF information, >50% missing values, or failed feature extraction were excluded. K-means clustering identified three circadian phenotypes: restorative (A, B), characterized by nocturnal decline with early troughs; riser (C, D), marked by blunted or reversed rhythms with daytime troughs; and irregular (E, F), showing attenuated or inconsistent rhythmicity. Mean trajectories with cosine curve fitting (red lines) highlight trough timing for each cluster. Consistent clustering across both registries supports the reproducibility of AI-ECG-derived circadian phenotypes.

### Characterization and Temporal Reproducibility of Circadian AI-ECG Phenotypes

To translate the unsupervised clusters into a clinically interpretable metric, we derived a binary phenotype based on the day–night AI-ECG age difference (ΔAge). Using a minimum-distance classifier, participants were categorized as Restorative (nocturnal decline; ΔAge >0.6 years) or Disrupted (blunted decline or paradoxical nocturnal increase; ΔAge ≤0.6 years). These classifications captured distinct circadian trajectories (**Figure 3**): the Restorative group exhibited a physiological dipping pattern with a nadir near midnight, whereas the Disrupted group displayed a non-dipping or rising pattern. The prevalence of the Restorative phenotype was similar across cohorts (47.6% in S-Patch, 50.2% in Memo Patch; **Table 2**). This phenotype demonstrated moderate reproducibility, with short-term intra-individual concordance of 76% (Cohen’s κ=0.518) and reproducibility over extended monitoring periods (ICC 0.51–0.54), supporting ΔAge as a reproducible circadian phenotype over consecutive days (**Table 3**).

**Figure 3.**
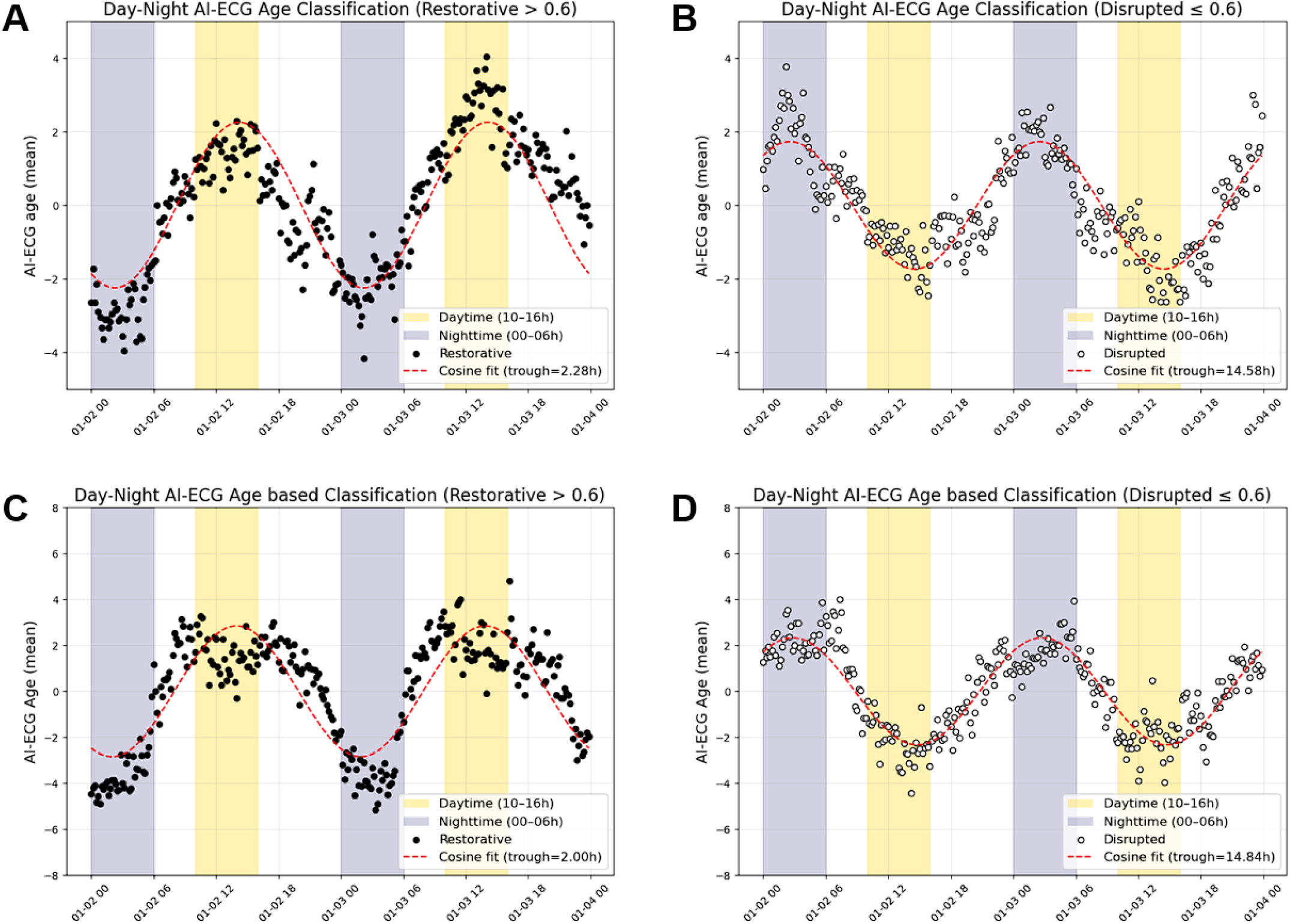
Day–Night AI-ECG Age Classification of Circadian Patterns. This figure illustrates a clinically parsimonious classification of circadian rhythmicity in AF-naive participants, based on the day–night AI-ECG age difference (ΔAge = mean daytime [10:00–16:00] − mean nighttime [00:00–06:00]). Using a threshold of ΔAge > 0.6 years, participants were categorized as having a Restorative pattern (A, B), characterized by a clear nocturnal decline and consistent troughs near midnight. Those with ΔAge ≤ 0.6 years were classified as Disrupted (C, D), exhibiting blunted or absent nocturnal dipping with flattened circadian profiles.

**Table 2.**
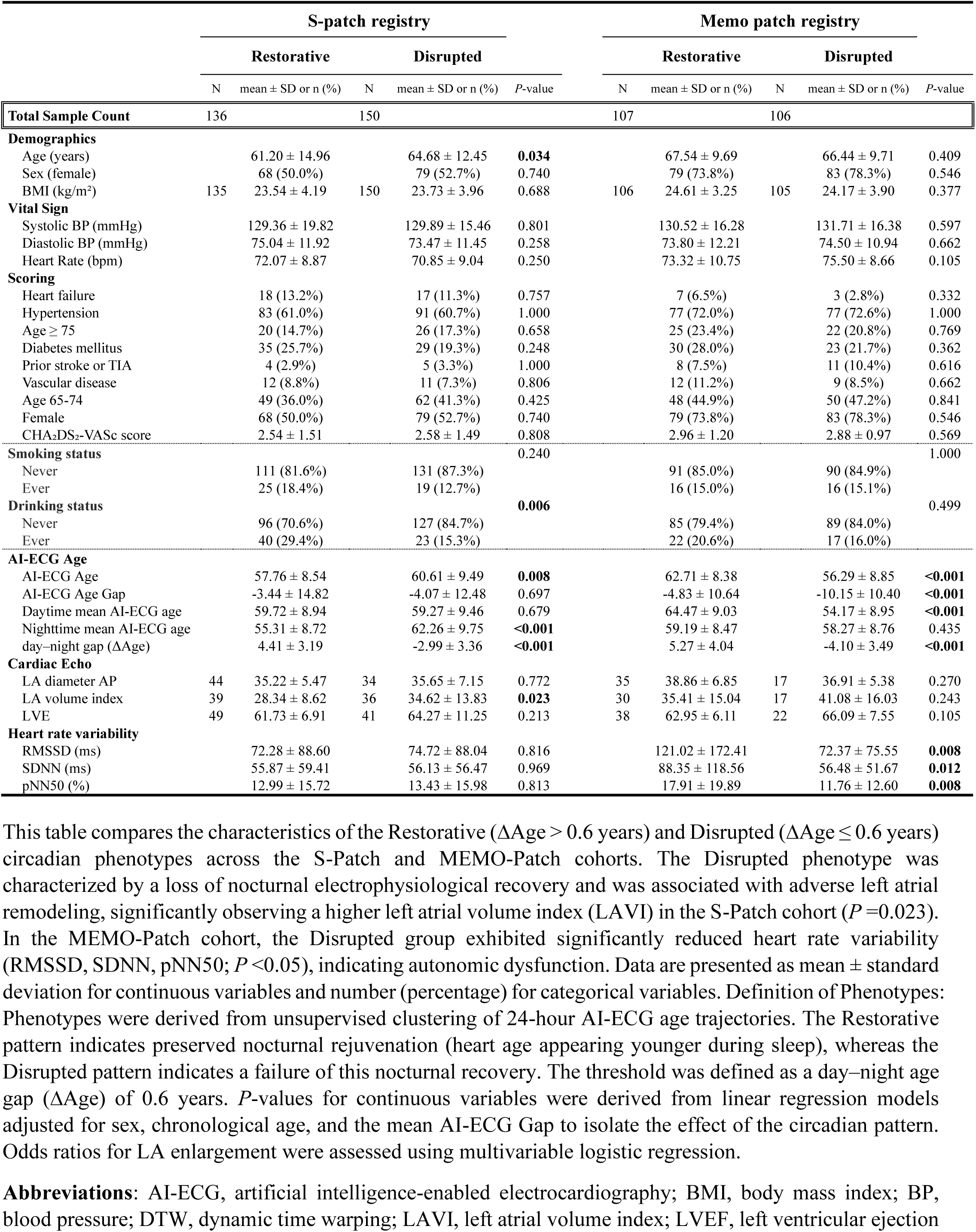
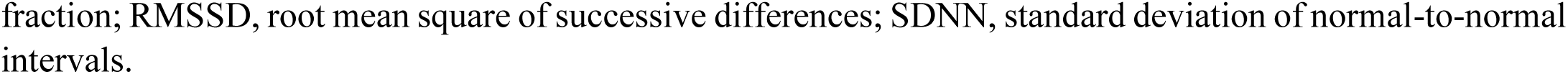
Clinical, Echocardiographic, and Heart Rate Variability Characteristics Stratified by Circadian AI-ECG Phenotypes.

**Table 3.** Assessment of Short- and Long-Term Consistency of AI-ECG Circadian Phenotypes.

|  |  |  |  |
| --- | --- | --- | --- |
| **A. Short-term Consistency** |  |  |  |
| **Contingency Table** | **Period 1 Restorative** | **Period 1 Disrupted** | **Total** |
| Period 2 Restorative | 86 | 30 | 116 |
| Period 2 Disrupted | 21 | 75 | 96 |
| *Total* | *107* | *105* | *212* |
| **Statistics** | **Value** | **95% CI / P-value** |  |
| Concordance Rate | 0.759 | - |  |
| Cohen's Kappa ($\kappa$) | 0.518 | - | |
| Cramer's V | 0.511 | P < 0.001 |  |
| Spearman's Correlation ($\rho$) | 0.520 | P < 0.001 | |
| **B. Long-term Consistency** | **6-day (Three)** | **8-day (Four)** |  |
| Percent Agreement | 0.654 | 0.550 |  |
| Fleiss' Kappa ($\kappa$) | 0.537 | 0.507 | |
| Pairwise $\kappa$ -mean | 0.538 | 0.507 | |
| ICC (Two-way mixed) | 0.539 | 0.509 |  |

This table summarizes the reproducibility of restorative and disrupted circadian phenotypes. Short-term consistency, evaluated between two sequential 48-hour periods, showed a concordance of 75.9% and moderate agreement (Cohen's $\kappa$ = 0.518; Cramer's V = 0.511, P < 0.001). Long-term consistency, assessed over 6-day and 8-day periods, demonstrated sustained stability with percent agreement ranging from 55% to 65% and ICCs between 0.51 and 0.54. Overall, the AI-ECG-derived day-night age gap phenotype exhibits moderate but consistent reproducibility across extended monitoring durations.
**Abbreviations:** AI-ECG, artificial intelligence-enabled electrocardiography; ICC, intraclass correlation coefficient; CI, confidence interval.

### Association with Left Atrial Remodeling and Structural Phenotypes

The Disrupted circadian phenotype was associated with adverse LA remodeling (**Figure 4**). In the S-Patch cohort, participants with a Disrupted phenotype exhibited higher LAVI compared with the Restorative group (34.6 ± 13.8 vs. 28.3 ± 8.6 mL/m²; p=0.023). This association persisted after adjustment for age, sex, BMI, and baseline AI-ECG age gap (adjusted mean difference: 5.64 mL/m²; 95% CI: 0.39–10.89, p=0.020). Furthermore, the Disrupted phenotype was associated with higher odds of severe LA enlargement (adjusted OR: 5.29; 95% CI: 1.45–19.27, p=0.012). Although the Memo Patch cohort showed a similar directionality without reaching statistical significance in isolation (41.08 ± 16.03 vs. 35.41 ± 15.04 mL/m²; p=0.243), a pooled meta-analysis of both cohorts confirmed the association. The Disrupted phenotype was associated with a pooled adjusted mean increase in LAVI of 6.09 mL/m² (95% CI: 1.46–10.72; I²=0%; p=0.010) and a 4.17-fold higher odds of severe LA enlargement (pooled adjusted OR: 4.17; 95% CI: 1.58–10.99; I²=0%; p=0.004).

**Figure 4.**
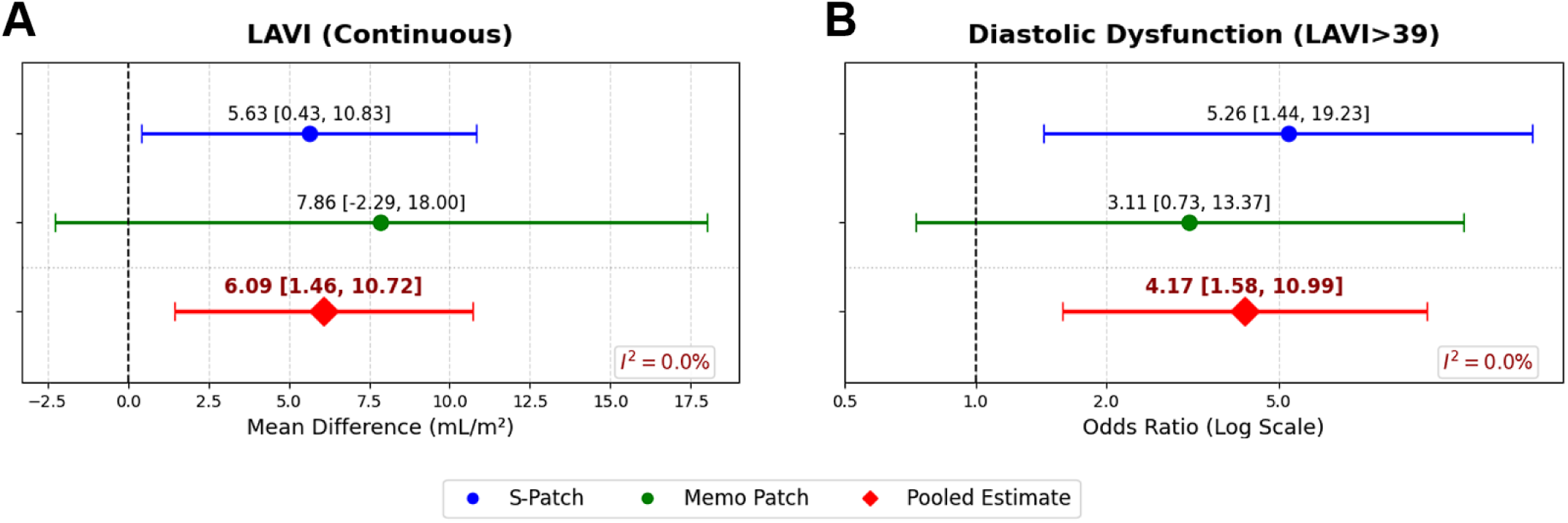
Association of Disrupted Circadian AI-ECG Phenotypes with Left Atrial Remodeling. Forest plots illustrate the meta-analysis of the S-Patch and MEMO-Patch cohorts, comparing restorative (ΔAge > 0.6 years) and disrupted (ΔAge ≤ 0.6 years) circadian phenotypes. (A) Participants with a disrupted phenotype exhibited a significantly larger left atrial volume index (LAVI) compared with the restorative group (Pooled Mean Difference: 6.09 mL/m²; 95% CI, 1.46–10.72; p = 0.010). (B) The disrupted phenotype was associated with four-fold higher odds of enlarged left atrium, defined as LAVI > 39 mL/m² (Pooled Odds Ratio: 4.17; 95% CI, 1.58–10.99; p = 0.004). Models were adjusted for age, sex, BMI, and mean AI-ECG gap. The pooled estimates (red diamonds) were calculated using a fixed-effects model, showing no heterogeneity across cohorts (I^2^ = 0.0%). **Abbreviations**: AI-ECG, artificial intelligence-enabled electrocardiography; CI, confidence interval; LAVI, left atrial volume index; BMI, body mass index.

Of note, sex-stratified analyses (**Figure S2**) suggested effect modification by sex: the association with LA remodeling was pronounced in females (pooled mean difference: 10.11 mL/m²; 95% CI: 4.09–16.13; p=0.001) but not observed in males (pooled mean difference: −0.75 mL/m²; 95% CI: −7.92–6.42; p=0.837).

### Independence from Conventional Autonomic and Static Metrics

While the overall circadian trajectories of HRV were qualitatively similar between groups (**Figure S3, S4**), differences in autonomic indices were observed by AI-ECG age phenotype. In the Memo Patch cohort, the Disrupted phenotype was associated with lower global HRV compared to the Restorative phenotype, including lower SDNN (56.48 ± 51.67 vs. 88.35 ± 118.56 ms; p=0.012), RMSSD (p=0.008), and pNN50 (p=0.008) (**Table 2**, **Figure S4**). A random-effects meta-analysis encompassing both cohorts showed a consistent direction toward lower HRV in the Disrupted group; however, these differences did not reach statistical significance in the overall population (**Figure S5**). Beyond autonomic parameters, the day–night AI-ECG age gap provided complementary information not captured by static AI-ECG age estimates. The Disrupted phenotype was not associated with the prevalence of traditional comorbidities—including hypertension, diabetes, and heart failure—nor with static AI-ECG metrics such as overall mean ECG age (**Figure S6**).

## Discussion

This study aimed to characterize the circadian dynamics of AI-ECG age in an AF-naïve population and to examine whether these patterns are associated with structural cardiac health. Using continuous single-lead wearable ECGs, we demonstrated three main findings. First, unsupervised machine learning identified consistent circadian trajectories of AI-ECG age, enabling classification into two phenotypes: Restorative (a nocturnal decline) and Disrupted (blunted decline or paradoxical nocturnal increase). Second, this binary phenotype showed moderate short-term agreement across consecutive days, suggesting that the day–night difference (ΔAge) captures a recurring individual trait rather than random fluctuation, while also indicating that within-person variability remains substantial and warrants further characterization. Third, and most importantly, the Disrupted phenotype was associated with adverse left atrial remodeling, including higher left atrial volume index (LAVI) and increased odds of severe LA enlargement.

This study leverages multi-center registries of continuous, wearable-derived AI-ECG age and captures high-resolution circadian dynamics of electrophysiological aging in free-living conditions. Prior work on circadian cardiovascular patterning has largely focused on arrhythmia-present populations (16,21) or autonomic indices such as HRV (15,22). To our knowledge, this is the first study to characterize circadian patterning of a wearable-derived digital biomarker reflecting overall electrophysiological aging in an AF-naïve population. Importantly, our morphology-based signal captures aspects of electrophysiological aging related to conduction and repolarization (5) that are not fully reflected by conventional rate-based measures.

Cardiovascular physiology is organized by circadian biology, and electrophysiological properties show reproducible day–night variation across molecular, cellular, and systems levels (12). In this context, attenuation of a nocturnal decline in AI-ECG age may reflect blunted diurnal modulation of electrophysiological features related to conduction and repolarization (5,23,24). Although the present study cannot establish mechanism, our central empirical observation is that an attenuated nocturnal decline in AI-ECG age is associated with LA enlargement—a marker of adverse atrial remodeling. This association was independent of conventional comorbidities and static AI-ECG age, suggesting that the circadian pattern captures a dimension of structural vulnerability not reflected by point-in-time assessments alone. Given that LA dilation is a recognized feature of atrial cardiomyopathy (25,26), diurnal AI-ECG age phenotyping may offer a non-invasive, wearable-derived window into early atrial substrate changes, though this interpretation requires prospective validation.

The concept that nocturnal physiological recovery is protective is well established in cardiovascular medicine. In blood pressure monitoring, a preserved nocturnal dipping pattern is associated with favorable outcomes, whereas non-dipping or riser patterns confer increased cardiovascular risk (13,14). Our findings extend this paradigm to electrophysiological aging: attenuation of a nocturnal decline in AI-ECG age—a morphology-based biomarker of overall electrophysiological health—was associated with adverse atrial remodeling. This pattern is conceptually consistent with recent device-based observations from the LOOP study, in which AF-naïve individuals undergoing ILR screening were clustered into circadian AF phenotypes; those with predominantly midnight–morning AF onset exhibited higher AF burden and greater disease progression compared with a daytime phenotype (16). Taken together, these findings suggest that individuals who fail to achieve nocturnal electrophysiological recovery—whether reflected by blunted dipping in blood pressure, attenuated decline in AI-ECG age, or increased nocturnal arrhythmia burden—may share a common vulnerability to adverse cardiovascular remodeling and outcomes. Importantly, our cohort was restricted to AF-naïve individuals with no AF detected during monitoring, positioning circadian AI-ECG age phenotyping as a potential upstream marker of this vulnerability, measurable before arrhythmia emerges. Prospective longitudinal studies are needed to determine whether the Disrupted phenotype predicts incident AF and whether it identifies individuals who, once AF develops, are more likely to follow an unfavorable trajectory.

Several aspects of these findings merit further discussion. First, the moderate reproducibility of the circadian phenotype (Cohen’s κ=0.518; ICC 0.51–0.54) should be interpreted in the context of broader cardiovascular phenotyping. In ambulatory blood pressure monitoring, even well-established clinical classifications such as masked and white-coat hypertension show limited long-term reproducibility, with only approximately 40% of patients maintaining the same classification across repeated measurements (27). That our wearable-derived phenotype achieves comparable stability from a 48-hour recording suggests it captures a recurring physiological trait, though within-person variability remains substantial and longer-term consistency should be evaluated in future studies. Second, the observed sex-specific pattern—stronger associations with LA remodeling in females and attenuation in males—warrants dedicated investigation. This heterogeneity may reflect differences in exposure profiles (e.g., smoking and alcohol), sleep and breathing disorders, and sex-specific atrial remodeling pathways, including hormonal modulation and differences in atrial fibrosis patterns (28). Future prospective studies should incorporate behavioral and sleep-related measures (e.g., actigraphy, obstructive sleep apnea screening), chronotype, and hormonal status to clarify effect modification and to evaluate whether monitoring circadian electrophysiological dynamics improves prediction of incident AF and related outcomes beyond established risk factors.

### Limitations

Several limitations should be acknowledged. First, single-lead AI-ECG age estimation entails information loss relative to 12-lead ECGs and may yield higher absolute error; however, our primary focus on within-person diurnal contrast (ΔAge) reduces dependence on absolute calibration and showed consistent associations across cohorts. Second, the cross-sectional observational design precludes causal inference, and residual confounding—particularly sleep quality, obstructive sleep apnea, physical activity timing, and chronotype—may influence phenotype assignment. Third, day–night windows were defined using fixed clock times and were not anchored to individualized sleep or circadian phase (e.g., actigraphy or sleep staging), which may introduce nondifferential misclassification that would be expected to bias associations toward the null. Fourth, echocardiographic validation was available in a modest subset (n=122); although associations were statistically significant and supported by meta-analysis with minimal heterogeneity, findings should be interpreted as hypothesis-generating. Fifth, the cohorts were predominantly East Asian, limiting generalizability. Prospective longitudinal studies across diverse populations are needed to confirm whether the Disrupted phenotype independently predicts incident AF, stroke, or heart failure.

## Conclusions

Wearable-derived AI-ECG age exhibits circadian patterns in AF-naïve individuals, with unsupervised learning identifying distinct trajectories. Attenuation of a nocturnal decline—the Disrupted phenotype—is associated with left atrial enlargement, independent of conventional comorbidities and static AI-ECG age metrics. These findings suggest that circadian electrophysiological aging phenotyping may capture a dimension of atrial structural vulnerability not reflected by point-in-time assessments, and support prospective studies to evaluate its clinical utility.

## Declarations

## Acknowledgements

We express our gratitude to Severance Hospital for providing invaluable ECG data and Daeun Joung. S.H.P. acknowledges support from the Yonsei University College of Medicine MSTP Scholarship.

## Sources of Funding

This study was funded by funded by the Ministry of Health & Welfare, Republic of Korea (RS-2022-KH125397; RS-2022-KH129902, RS-2023-00265440, RS-2024-00397290, HI22C0452), the Ministry of Science and ICT, Republic of Korea (RS-2025-24533659), Samjin, Yuhan, Wellysis, and HUINNO Corporation.

## Disclosures

S.H.P. declares no competing interests beyond the institutional funding reported above. J.H.J., D.L., and H.T.Y. declare no competing interests. J.K. is a shareholder of Wellysis Corp. and reports pending patent applications related to atrial fibrillation prediction using AI (United States Application No. 18/636,402, filed 15 April 2024; Republic of Korea Application No. 10-2023-0069397, filed 30 May 2023). D.K. and J.J. are shareholders of HUINNO Corp. S.C.Y. serves as the Chief Executive Officer of PHI Digital Healthcare, reports grants from Daiichi Sankyo, and is a coinventor of granted Korean Patents (DP-2023-1223, DP-2023-0920) and pending Patent Applications (DP-2024-0909, DP-2024-0908, DP-2022-1658, DP-2022-1478, DP-2022-1365, PATENT-2025-0039190, PATENT-2025-0039191, PATENT-2025-0039192, PATENT-2025-0039193, PATENT-2025-0039194), all unrelated to the present work. B.J. has served as a speaker for Bayer, BMS/Pfizer, Medtronic, and Daiichi-Sankyo, and received research funds from Samjin, Yuhan, Medtronic, Boston Scientific, and Abbott Korea.

## Author Contributions

S.H.P. and S.C.Y. co-conceived and led the study. S.H.P. served as the primary author, developed the analytical framework, implemented the AI model pipeline, performed all primary statistical analyses, and drafted the manuscript including tables and figures. S.C.Y. directed the overall research, established the statistical analysis plan, and provided critical scientific supervision. B.J., as a co-supervising professor, designed and directed the prospective cohort registries, facilitated clinical data acquisition, and provided senior clinical oversight. J.H.J. supported the statistical analyses and provided research assistance. H.T.Y. participated in the review of the manuscript. J.K., D.L., D.K., and J.J. (industry collaborators) secured, processed, and provided the single-lead wearable ECG data, and supported technical coordination. All authors critically reviewed and approved the final manuscript.

## Notes

### Clinical Trial

NCT05119725, NCT05355948

### Author Declarations

This study was performed in accordance with the ethical principles of the Declaration of Helsinki. For the prospective wearable validation cohorts, data were obtained from two pre-registered clinical trials approved by their respective Institutional Review Boards: The S-Patch registry (IRB No. 1-2021-0002; https://ClinicalTrials.gov identifier: NCT05119725, registered November 2021) and the Memo Patch registry (IRB No. 1-2022-0008; https://ClinicalTrials.gov identifier: NCT05355948, registered May 2022) Informed consent was obtained from all participants in these prospective registries. All datasets utilized in this study-specifically those derived from the S-Patch and Memo Patch registries-were fully de-identified and anonymized prior to use by the research team. The data collection and processing were conducted in accordance with the protocols approved by the Institutional Review Board of Severance Hospital (IRB No. 1-2021-0002 and 1-2022-0008).

